# Effects of supervised exercises on pain and disability in patients with hip or knee osteoarthritis: A systematic review and meta-analysis

**DOI:** 10.1101/2023.02.09.23285694

**Authors:** Jean Mapinduzi, Gérard Ndacayisaba, Penielle Mahutchegnon Mitchaϊ, Bruno Bonnechère, Kossi Oyéné

## Abstract

**Background:** Hip and knee osteoarthritis are among the most prevalent and disabling conditions affecting mainly older adults, with a risk of undergoing a total hip or knee replacement for the end stage. Increasing recommendations of conservative treatments have been reported as the first-line strategy for the initial management of this condition.

**Objective:** This review sought to investigate the effects of supervised exercises versus non-pharmacological conservative therapies to reduce pain and disability levels on patients with hip or knee osteoarthritis.

**Methods:** Three databases (PubMed, Cochrane Library, and ScienceDirect) were systematically searched for randomized-controlled trials published between 01-01-2001 and 31-10-2022. PEDro scale was used to assess the methodological quality of the included studies. The PRISMA guidelines were applied for this review.

**Results:** Twelve randomized-controlled trials of moderate-to-high quality were included in this review. The studies involved 1,049 participants with a mean age (SD) of 64 (6) years old. The duration of the intervention and follow-up varied from 2 weeks to 16 months. Supervised exercises for hip or knee osteoarthritis were significantly less effective in terms of pain reduction (SMD=-0.40 [95%CI 0.16, 0.65], p=0.001) compared to home-based exercises (active musculoskeletal therapies), but not in terms of disability reduction (SMD=-0.04[95%CI −0.43, 0.36], p=0.86). There was a non-significant difference of supervised exercises compared to passive musculoskeletal therapies in terms of disability (SMD=0.21[95%CI −0.09, 0.50], p=0.17), or pain reduction (SMD=-0.19; [95%CI −0.57, 0.19], p=0.33).

**Conclusion:** Supervised exercises were found to be less effective in reducing pain, but not in disability reduction when compared to home-based exercises.

**Systematic review registration:** Prospero CRD42021271912

## 1. Introduction

Osteoarthritis (OA) is among the most prevalent and disabling conditions affecting mainly the older adults, with an estimated 25-40% lifetime risk of symptomatic OA in people who live to age 85 [1, 2]. Almost 10% lifetime risk of undergoing a total hip or knee replacement for end-stage of OA [1, 2]. It has been reported that OA affects more than 240 million people worldwide and is the most frequent reason for activity limitations in adults [3, 4]. The knee and hip joints are the most affected weight bearing joints [5]. The characteristics of OA are commonly structural and functional failure of the articular and abarticular elements [6]. Radiologically, joint space narrowing, bony sclerosis, osteophyte formation and deformity of articular surfaces are mostly evident in OA condition in nearly 30% of people older than 45 years, women being more affected than men in both conditions [6]. Moreover, decreased joint range of motion, altered gait patterns, reduced lower limb proprioception and balance, surrounding muscle weakness, crepitus, occasional effusion, variable degrees of inflammation, worsening of the general health status and quality of life are the main clinical presentation [3, 7].

This physical condition can lead to poor physical function, fatigue related to sleep disturbance, reduced work productivity (through absenteeism), and increased direct and indirect health care costs [8]. Hip and knee OA is commonly associated with comorbidities, which may stem from lack of physical activity, medication toxicity, and the effects of inflammatory cytokines, and this may lead to a 20% higher age-adjusted mortality [3].

Many risk factors have been reported in the literature. On one side there are those at the joint level, such as abnormal joint morphology, developmental dysplasia of the joint, weakness in stabilizing muscles of the joint, joint injury and labral tears, etc. These can lead to the damage of the underlying cartilage via its response to shear stress [9]. On the other side, there are the whole person level risk factors, such as age, sex, weight, genetics, ethnicity, occupation (high-impact physical activity or sport), and diet [9].

Increasing recommendations of non-pharmacological treatments in the literature have been reported as a first-line strategy for the initial management of OA [10]. It has been suggested that non-operative management of hip or knee OA can prevent, delay the impact of disability, or postpone operative management. Interventions such as patient education, manual therapy, exercise modalities (including supervised exercises) have been recommended as the first choice of treatment [11].

Previous systematic reviews have summarized available evidence to investigate the effects of manual and exercise therapies in reducing pain and disability in patients with hip or knee OA [12–14]. However, the lack of sufficient number of hip or knee OA specific randomized clinical trials (RCTs) prevented authors from performing analysis at the time. Moreover, to our knowledge, no recent systematic review with meta-analysis has been conducted to explore the effectiveness of supervised exercises on pain and disability in hip or knee OA.

The aim of this study was to investigate the effects of supervised exercises versus non-pharmacological conservative therapies in reducing pain and disability in patients with hip or knee OA.

## 2. Methods

The review protocol was registered into PROSPERO under the registration number CRD42021271912. The Preferred Reporting Items for Systematic Reviews and Meta-Analyses (PRISMA) guidelines were employed for conducting this review [15].

### 2.1 Search Strategy

PubMed, Cochrane Library, and ScienceDirect databases were systematically searched for relevant RCTs published between 01-01-2001 and 31-10-2022.

The period from 2001 was considered because it corresponds with the introduction and implementation of the International Classification of Functioning, disability, and health (ICF) as the international standard to describe and measure health and disability.

The studies, along with their titles, abstracts, and data, were identified based on these core concepts, (((((((((((((“osteoarthritis, hip”[MeSH Terms]) OR “osteoarthritis”) OR “hip osteoarthritis”) OR “knee osteoarthritis”) OR “coxarthrosis”) OR “gonarthrosis”) AND “aerobic” AND “exercis*”[MeSH Terms]) OR “anaerobic exercis*”) OR “supervised exercis*”) OR “supervised physical exercis*”) OR “therapeutic exercis*”), and screened by two independent reviewers (MJ and NG). Only eligible full-text studies were retrieved and then screened again by the same reviewers. In addition, the reference lists of the identified studies were manually checked for further inclusions.

### 2.2 Eligibility criteria

The following PICO’s criteria were defined to guide our search strategy:

#### Participants/population

Patients (men and women) suffering from hip or knee OA diagnosed using X-ray.

#### Interventions

These were supervised exercises, such as hip and knee flexibility or stretching and strengthening exercises, pelvic-tilt exercises with pelvic floor muscle (PFM) voluntary contraction, as well as exercises involving hip flexion, extension, abduction, knee flexion, extension combined or not with knee rotation exercises, and functional exercises. These exercises were supervised by a physiotherapist in hospital or rehabilitation center services.

#### Comparators

These were other non-pharmacological conservative therapies, such as active musculoskeletal therapies (Home-based exercises), or passive musculoskeletal therapies (MSKTs) (massage, electrotherapy, acupuncture, hot or cold packs) that were used as controls.

#### Outcome measures

Pain and disability levels.

#### Study design

RCTs written in English or French.

Search results were stored and organized using EndNote 20 computer software. The flowchart of the study selection is presented in Figure 1.

**Figure 1:**
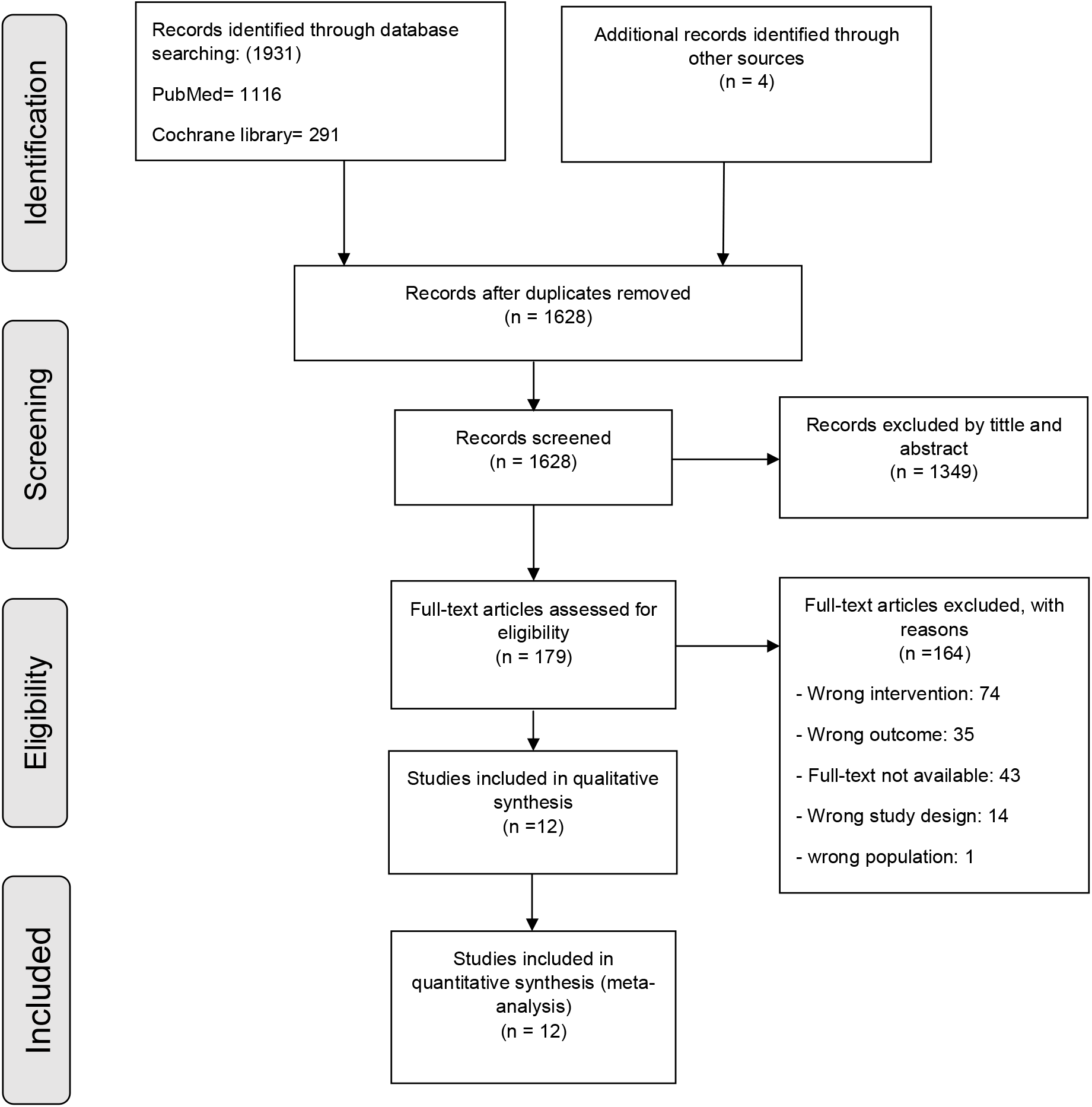
Flowchart of the study selection.

### 2.3 Methodological assessment of RCTs

The PEDro scale, which is deemed a valid and reliable tool for assessing RCTs [16, 17], was used for methodological quality assessment. RCTs’ quality was blindly judged by two different reviewers (MJ and NG) to minimize potential bias [18]. The final decision about each RCT quality was made by reaching a consensus. In case of discordance, a third collaborator (KO) was consulted. Accordingly, RCTs were divided into three categories, as follows: low quality (0-3 / 10), moderate quality (4-6 / 10), and high quality (7-10 / 10) [18].

### 2.4 Data extraction

The following information was extracted from the included studies: characteristics of patients (age, sex, BMI, and OA location), number of participants, type of exercise interventions, intervention durations and follow-ups, main outcome measures, and results. This information was collected, classified, summarized, and displayed in Table 1 by the same reviewers (MJ and NG).

**Table 1:** The Characteristics of the included studies.

| Study | Country | PEDro score | Purpose | Site | Participants | Interventions | Number of participants | Duration & FU* period | Outcome measures | Main results |
| --- | --- | --- | --- | --- | --- | --- | --- | --- | --- | --- |
| Tunay et al., 2010 | Turkey | 5/10 | To establish the effects of hospital and home-based proprioceptive and strengthening exercise programs on proprioception, pain, and functional status in patients with knee OA* | Knee | N= 60<br>Age<br>G1= 50.2±9.1<br>G2= 54.4±8<br>BMI*<br>G1 = 27.5±4.3<br>G2= 28.8±5.1 | G1: hospital-based proprioceptive and strengthening exercises<br>G2= home-based proprioceptive and strengthening exercises | G1; n = 30<br>G2; n = 30 | 5 times/ week<br>6 weeks (30 sessions) | WOMAC*<br>VAS*<br>MFSS*<br>TUG * | Both hospital and home-based exercise programs decreased joint symptoms and improved function in patients with knee OA*. |
| Ihana et al., 2017 | Brazil | 6/10 | To investigate the effects of therapeutic exercises associated with the pompage technique on pain, balance and muscular strength in a group of elderly women with a knee OA | Knee | N = 22(females)<br>Age:<br>G1: 65±4;<br>G2: 67±6<br>BMI*:<br>G1: 30±4;<br>G2: 29±5 | G1: supervised exercises<br>G2: Patient education | G1; n = 11<br>G2; n = 11 | 12 Weeks | WOMAC<br>OSI* | Supervised exercises showed better results for pain, balance and muscle strength compared to patient education after 12 weeks. |
| Marconcin et al., 2017 | Portugal | 6/10 | To assess the effectiveness of a 12-week self-management and exercise intervention in elderly individuals with knee OA. | Knee | N = 67 (47 females)<br>Age:<br>G1: 70±6;<br>G2: 68±5<br>BMI*<br>G1: 30±5;<br>G2: 32±5 | G1: patient education<br>G2: supervised exercises | G1; n = 32<br>G2; n = 35 | 12 weeks | KOOS*;<br>EQ-5D-5L* VAS*;<br>6MWT* (m) | It was found important to combine self-management and exercises for improving pain and disability in knee OA |
| Tuğba et al., 2016 | Turkey | 6/10 | To compare the effects of low-intensity exercise programs for lower extremities, either supervised or at home, on pain, muscle strength, balance and the | Knee | N = 56 (39 females)<br>Age:<br>G1: 60 (49–84);<br>G2: 59 (51–80)<br>BMI:<br>G1: 32±6; | G1: Supervised exercises<br>G2: HBE* | G1; n = 33<br>G2; n = 23 | 6 weeks | VAS, mm;<br>6-MWT, m;<br>Balanced score | Low-intensity supervised exercises were more effective than HBE in reducing post-activity pain levels and improving quadriceps and hamstring muscle strength. Both |
|  |  |  | hemodynamic parameters of knee OA patients. |  | G2: 30±4 |  |  |  |  | interventions were seen to be effective in improving rest pain and 6 MW distance in knee OA. |
| Kudo et al., 2013 | Japan | 4/10 | To evaluate effects of the mode of treatment delivery on the improvement of symptoms in knee OA, and to analyze potential risk factors affecting improvement after exercise therapies. | Knee | N = 209 (females)<br>Age:<br>G1: 64±6;<br>G2: 66±6<br>BMI:<br>G1: 24±3;<br>G2: 24±3 | G1: Supervised exercises<br>G2: HBE | G1; n = 81<br>G2; n = 128 | 12weeks | Normalized WOMAC | A significant improvement in WOMAC was observed in supervised exercises compared with HBE. |
| Hawkins et al., 2012 | England | 7/10 | To assess the feasibility of conducting a full-scale-randomized controlled trial investigating whether the addition of a 4-week supervised exercise class was more beneficial in reducing pain and improving function than HBE alone in treating patients following a corticosteroid injection for knee OA | Knee | N = 32 (18 females)<br>Age:<br>G1: 58±11<br>G2: 63±7 | G1: HBE<br>G2: Supervised exercises | G1; n = 15<br>G2; n = 17 | 2, 6, 12 weeks | WOMAC | Both groups had reduced pain and disability at week 12, with the supervised group demonstrating better outcomes than HEP alone. |
| Alagesan et al., 2011 | India | 7/10 | To determine the effectiveness of supervised exercise versus HBE for knee OA | Knee | N= 60 (35 females)<br>Age:<br>G1:50±3;<br>G2:49±3 | G1: supervised exercises<br>G2: HBE | G1; n = 30<br>G2; n = 30 | 2 months | NPRS* WOMAC | A significant decrease in pain and disability in favor of supervised exercises was found; both supervised and HBE being effective in treatment of knee OA. |
| Williamson et al., 2007 | England | 8/10 | To evaluate the effects of standardized western acupuncture and physiotherapy on pain and functional ability in patients with severe knee OA pain awaiting knee arthroplasty | Knee | N = 181<br>Age:<br>G1: 72±8;<br>G2: 70±9;<br>G3: 70±10<br>BMI:<br>G1: 31±6<br>G2: 33±6<br>G3: 33±6 | G1: Acupuncture<br>G2: Physiotherapy<br>G3: Control | G1; n = 60<br>G2; n = 60<br>G3; n = 61 | 6, 7, 12 weeks | OKS*;<br>50-m walk;<br>WOMAC score;<br>VAS | Patients with severe knee OA can achieve a short-term reduction in OKS when treated with acupuncture. Authors suggested the combination of both interventions in an out-patient group. |
| Bieler et al., 2016 | Denmark | 6/10 | To evaluate the effects of 4 months of supervised strength training; supervised Nordic Walking; and HBE on functional performance in persons with hip OA not on a waiting list for surgery. | Hip | N = 152 (103 females)<br>Age:<br>G1: 70±5<br>G2: 70±6<br>G3: 69±6<br>BMI:<br>G1: 27±5<br>G2: 28±5<br>G3: 27±5 | G1: Strength training<br>G2: Nordic walking<br>G3: HBE | G1; n = 50<br>G2; n = 50<br>G3; n = 52 | 16 weeks | WOMAC<br>SF-36* | Pain reduction in the NW group at 4 months was significantly greater compared with the HBE group. Discernable between-group differences were shown for improvements in SF-36 subscales in favor for the intervention groups compared with the HBE group. |
| Hermann et al., 2016 | Denmark | 7/10 | To investigate the efficacy and feasibility of progressive-type resistance training in hip OA scheduled for THA* | Hip | N=80 (52 females)<br>Age:<br>G1: 70±8; G2: 71±8<br>BMI:<br>G1: 28±5;<br>G2: 27±4 | G1: supervised exercises<br>G2: patient education + HBE | G1; n = 40<br>G2; n = 40 | 10 weeks | HOOS* | Progressive type resistance training was feasible, and resulted in significant improvement in self-reported outcomes and increased leg muscle power. |
| Fernandes et al., 2010 | Norway | 6/10 | To compare the efficacy of patient education and supervised exercise with that of patient alone for the management of pain in patients with hip OA | Hip | N = 109 (59 females)<br>Age:<br>G1: 57±10<br>G2: 58± 10<br>BMI:<br>G1: 24.9 ± 3.8<br>G2: 25±3 | G1: Patient education<br>G2: Patient education + supervised exercise | G1; n = 54<br>G2; n = 55 | 4, 10, 16 months | WOMAC | No significant difference in pain reduction over time was found between patient education + supervised exercises versus patient education alone; but adding supervised exercises to patient education may improve physical function. |
| Hoogeboom et al., 2010 | Netherlands | 8/10 | To evaluate the feasibility and preliminary effectiveness of therapeutic exercise before total hip replacement in frail elderly. | Hip | N = 21 (14 females)<br>Age:<br>G1: 77±3<br>G2: 75±5<br>BMI:<br>G1: 26±3<br>G2: 27±4 | G1: Supervised exercises<br>G2: Patient education | G1; n = 10<br>G2; n = 11 | 3 to 6 weeks | HOOS | Supervised exercises showed relevant preoperative improvements on the chair-rise time. |
| **Abbreviations :** BMI = Body Mass Index; EQ-5D-5L = EuroQoL five dimensions (Mobility, Self-care, Usual activities, Pain & discomfort, Anxiety & depression) ; FU = Follow-Up; HBE = Home-Based Exercises ; HOOS = Hip disability and Osteoarthritis Outcome Score ; KOOS = Knee injury and Osteoarthritis Outcome Score; MFSS= Monitorized Functional Squat System-Proprioceptive Test; 6MWT = 6-Minute Walk Test ; N = Number; NPRS = Numerical Pain Rating Scale; NW = Nordic Walking; OA = Osteoarthritis; OKS = Oxford Knee Score; OSI = Overall Stability Index; SF-36 = Short Form 36 questionnaire; TUG = Timed Up and Go test; THA = Total Hip Arthroplasty; VAS = Visual Analogue Scale; WOMAC = Western Ontario and McMaster Universities Osteoarthritis Index; |  |  |  |  |  |  |  |  |  |  |

### 2.5 Data synthesis and analysis

Results from studies exhibiting similar PICO’s (Patient, Intervention, Comparator, Outcomes, and study design) were considered for being pooled into separate meta-analyses. Pooled standard mean differences (SMDs) were calculated using Review Manager (RevMan V.5.4.1). Analyses being performed with random effects model, pooled estimates were calculated with their 95% confidence intervals (CIs) and an alpha level set at < 0.05 [19]. The effect sizes (ES) calculated with SMD were interpreted using Cohen’s method; the effect was defined as small (0-0.20), medium (0.20-0.50), or large (0.50-0.80) [20].

The efficacy of exercises was judged based on the SMD interpretation [21]. An SMD of zero means that the treatment in the intervention group (IG) and that in the control group (CG) display equivalent effects. If the improvement is associated with higher scores on the outcome measure, an SMD greater than zero indicates the degree to which the IG treatment is more effective than that administered to the CG, while an SMD less than zero indicates the opposite. If the improvement is associated with lower scores on the outcome measure, an SMD less than zero indicates the degree to which the IG treatment is more effective than that administered to the CG, while an SMD greater than zero indicates the opposite. In this review, considering the outcome measures, when SMD is less than zero, the improvement is in favor of the IG, while SMD greater than zero indicates the opposite.

A quantitative analysis was performed for meta-analysis. I^2^ was used as a statistical testing set to quantify inconsistency among studies [19]. This index describes the percentage of the variability in effect estimates that is due to heterogeneity. Thresholds for the I^2^ index interpretation can, however, be misleading, given that the relevance of the inconsistency may depend on several factors.

## 3. Results

### 3.1 Patients and study characteristics

Twelve RCTs were included and represented 1,049 participants with a mean age of 64 (6) years, and a mean BMI of 28.2 (4.5) (note that 2 studies [22, 23] did not report BMI). Eight RCTs [22–29] considered both genders with a majority of women, while 2 studies [30, 31] involved only women, and the remaining 2 others did not report patients’ gender [32, 33]. The sociodemographic characteristics of patients and studies have been presented in Table 1.

The PEDro score of the included studies ranged from 4 to 8/10. Five RCTs [22, 23, 26, 27, 32] were considered as being of high quality (PEDro score: 7 - 8/10), with the remaining ones [24, 25, 28–30, 33, 34] being of moderate quality (PEDro score: 4 - 6/10) (Table 2).

**Table 2:**
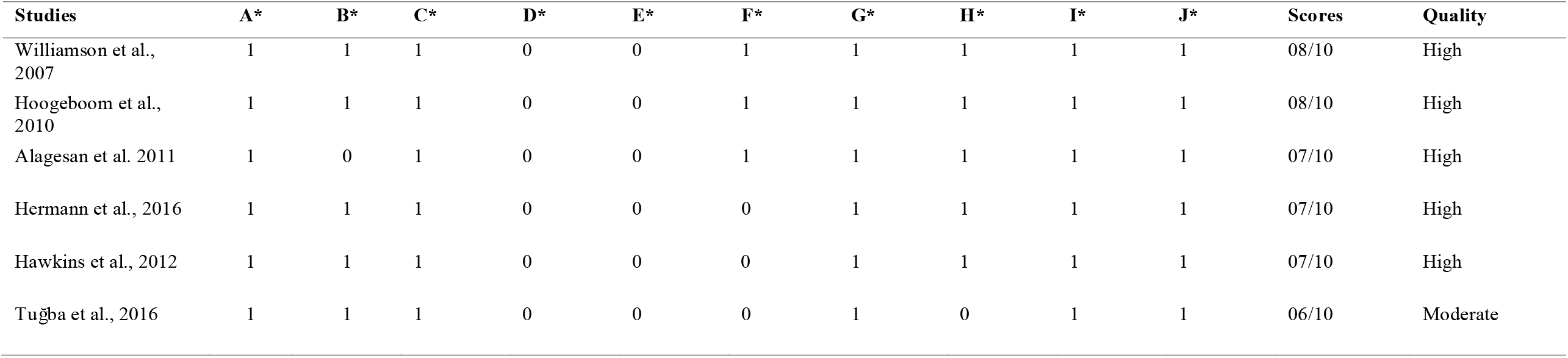
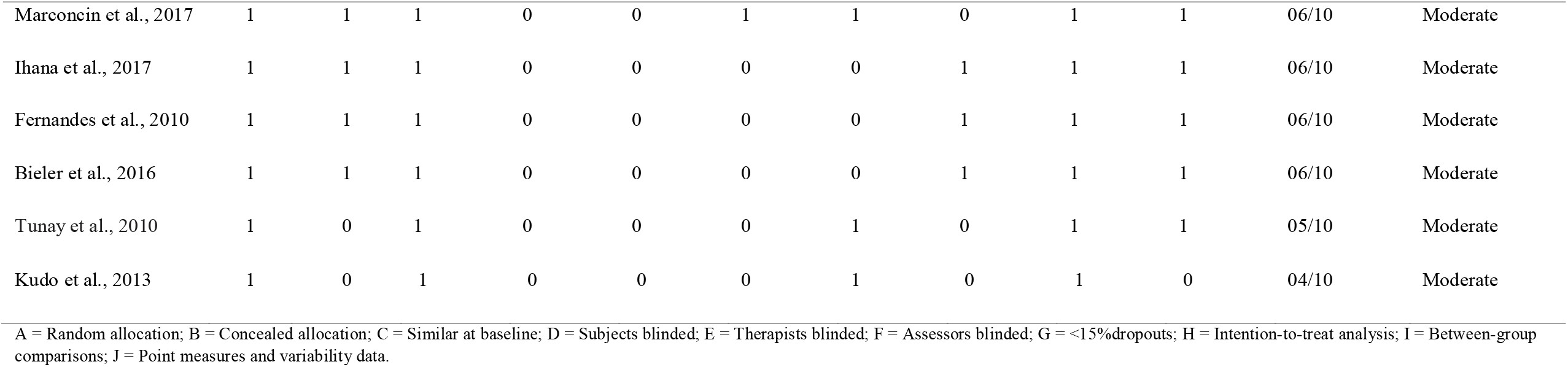
PEDro score of the included studies.

Different tests and scales were used in the included studies. For pain assessment, the Visual Analogue Scale was used in 3 RCTs [28, 32, 33] while the numerical pain rating scale was used in one RCT [22], but the assessment method was not further described in any study. Furthermore, 2 studies [26, 27] used the hip disability and osteoarthritis outcome score (HOOS) pain subscale, two others [29, 35] used the knee injury and osteoarthritis outcome score (KOOS) pain subscale, and others [23–25, 30, 32, 34] used the Western Ontario and McMaster Universities Osteoarthritis Index (WOMAC) pain subscale.

For self-reported function, WOMAC physical function subscale was used in 7 studies [22–25, 32–34], HOOS in two studies [26, 27], and KOOS was also used in 1 study [29]. In addition, the 6-minute walk test (6MWT) [28, 29], the Oxford knee score (OKS) and 50-m walk [32], and the Global Stability Index (OSI) [30] were used (Table 1).

Eight out of twelve (67%) studies [22, 23, 28–30, 32–34] involved patients with knee OA.

### 3.2 Description of the intervention exercises

All the included studies implemented a supervised exercise training program, either compared to active MSKTs (Home-based exercises) [22–24, 26, 28, 33, 34], or passive MSKTs (massage, electrotherapy, acupuncture, hot or cold packs) [25, 27, 29, 30, 32]. In the first part of this analysis, we are going to give the content of the program and then talk about its clinical efficacy.

#### 3.2.1 Supervised exercises versus home-based exercises (HBE)

The efficacy of supervised exercises versus HBE program was evaluated in 7 studies [22–24, 26, 28, 33, 34]. The number of participants in each study group varied from 15 to 128 participants, with a mean age of 62 (5) years. Five studies included both men and women, another one [34] only included women, while the remaining one [33] did not report patients’ gender. A variety of exercises was described with strengthening exercises to the muscles around the knee, the trunk, the hip, and the ankle. Additionally, proprioceptive training exercises, static bike, stretches, and open and closed-kinetic (squats and a calf raise, etc., if pain-free) chains and stabilization or neuromuscular control exercises (focusing on the knee, hip, pelvic, and trunk areas) were also described. These exercises were supervised by a physiotherapist in the intervention group, whereas the control group performed them at home. The total duration of the exercise programs varied from 6 to 16 weeks, with a frequency ranging from 1 to 6 times per week. The session duration varied from 40 min to 60 min per day, except for 2 studies [22, 33] which did not report this parameter.

#### 3.2.2 Supervised exercises versus passive MSKTs

The efficacy of supervised exercises versus passive MSKTs program was assessed in 5 studies [25, 27, 29, 30, 32]. The number of participants in each exercise group varied from 10 to 61 participants, with a mean age of 68 (7) years. Three studies [25, 27, 29] included both genders, while one study included only women [30]. The remaining one failed to report gender characteristics [32]. Supervised exercises consisted of a warm-up, leg-press through the full possible ranges of motion, bicycle ergometer, straight leg raises, calf stretches, TheraBand resisted knee extensions, wobble board balance training, knee flexion/extension sitting on a gym ball and freestanding peddle revolutions, static and dynamic balance and strength exercises for knee extensor and flexor muscles associated with knee pompage (patient in the supine position at the stretcher edge and the hanging leg placed between therapist’s legs. Three decompression maneuvers of 15 to 20s each were performed by a slight retreat from the therapist’s body [30]), functional exercises and flexibility exercises, functional physical activities of the patient’s daily living (stair climbing, sit to stand, step up and down). The total duration of the exercise programs varied from 6 to 16 weeks, the frequency ranging from once per day to 3 times per week. The control group received patient education and acupuncture.

### 3.3 Clinical efficacy

To quantify the efficacy of supervised exercises on pain and disability levels we then performed a meta-analysis to compare the effect of supervised exercises versus HBE or passive MSKTs.

#### 3.3.1 Effects of supervised exercises versus HBE

Five studies reported outcomes related to pain when comparing supervised exercises versus HBE [22, 23, 26, 28, 33]. We observed a statistically significant difference between the 2 interventions in terms of pain reduction (SMD=-0.40 [95%CI 0.16, 0.65], p=0.001) in favor of HBE.

Concerning disability level, 6 studies were included in our meta-analysis [23, 24, 26, 28, 33, 34]. No statistically significant difference was found between supervised exercises and HBE (SMD=-0.04[95%CI −0.43, 0.36], p=0.86). Individuals’ results are presented in Figure 2.

**Figure 2:**
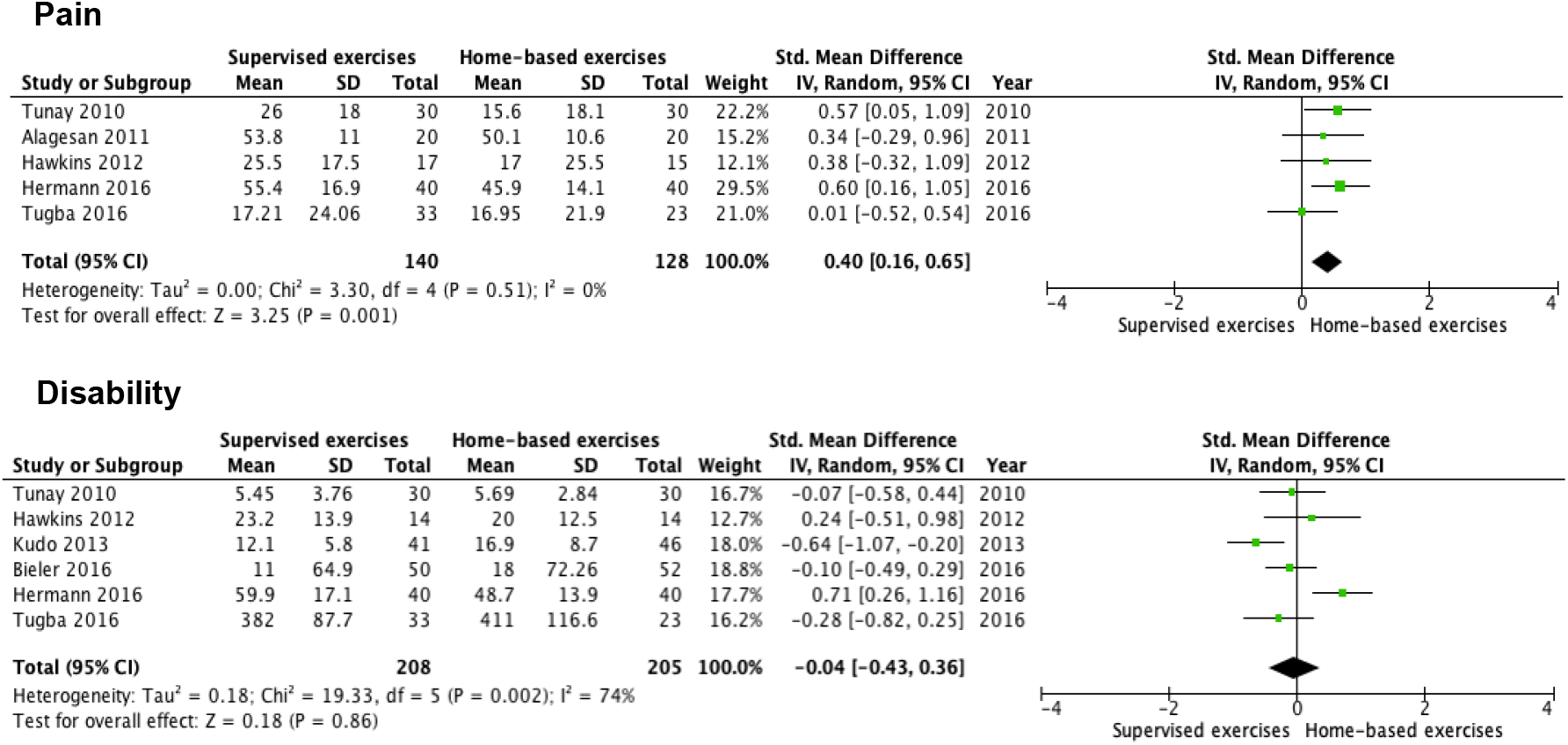
Effects of supervised exercises versus home-based exercises on pain and disability.

#### 3.3.2 Effects of supervised exercises versus passive MSKTs

Concerning the difference between supervised exercises and passive MSKTs, 4 studies were included for pain management [25, 27, 30, 32] and 5 for disability [25, 27, 29, 30, 32]. We did not find any significant difference in terms of pain reduction (SMD=-0.19; [95%CI −0.57, 0.19], p=0.33), nor for disability level (SMD=0.21[95%CI −0.09, 0.50], p=0.17), between the two interventions. Individuals’ results are presented in Figure 3.

**Figure 3:**
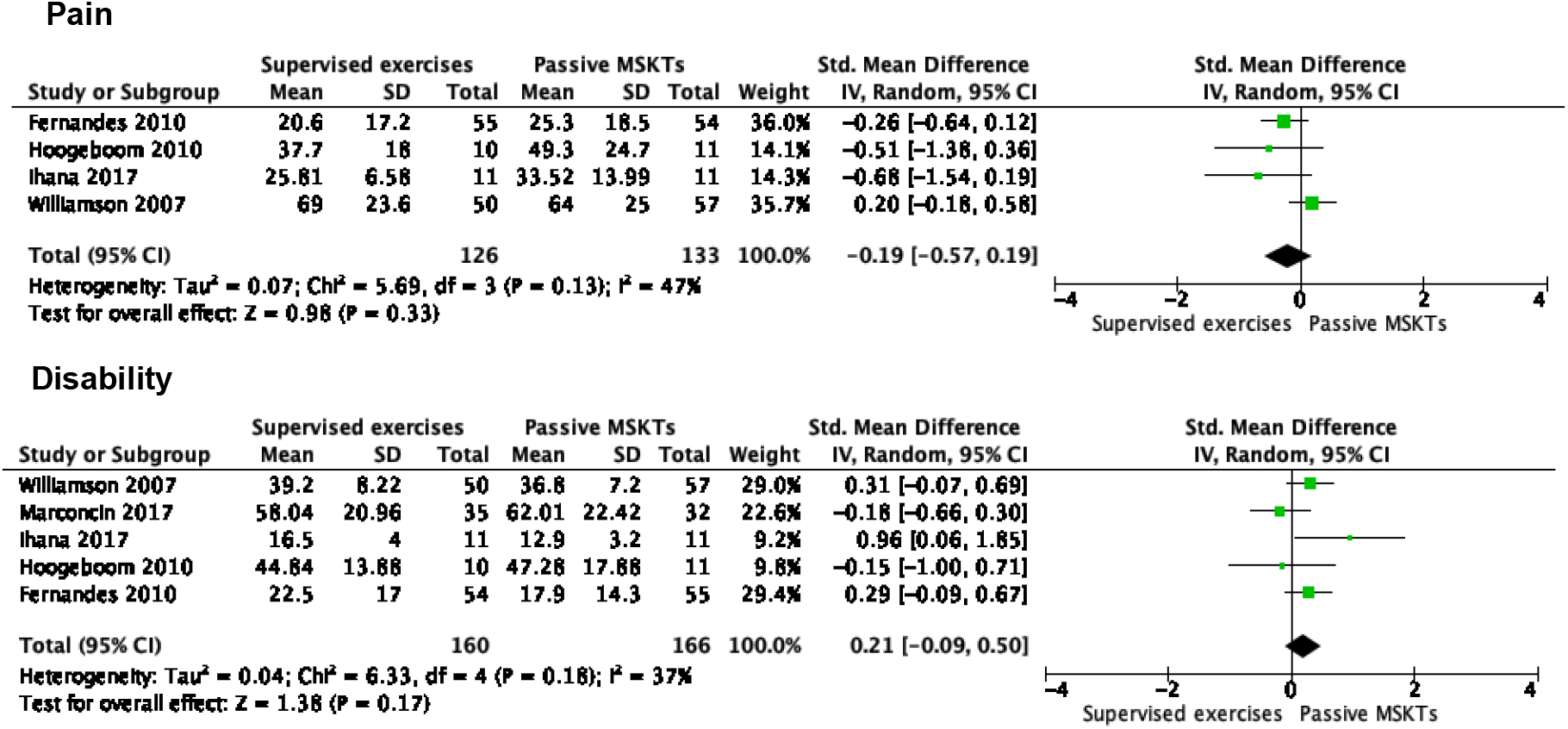
Effects of supervised exercises versus passive musculoskeletal therapies on pain and disability.

## 4. Discussion

This review aimed to investigate the effects of supervised exercises versus other MSKTs of two clinically-related outcomes; i.e., pain and disability, in patients with hip or knee OA. Subgroup analyses of 12 RCTs with 1,049 participants with radiographically confirmed symptomatic hip or knee OA showed, on one hand, a significant improvement in pain reduction, but not in disability, in favor of HBE. On the other hand, when comparing to passive MSKTs, no statistically significant differences were found between the two groups in terms of pain or disability reduction.

Concerning patient characteristics, most of the studies were about knee OA (67%). On one hand, this can be easily explained by the fact that the weight-bearing joint mechanical overload has been proposed as a possible mechanism to explain how weight increases the risk of knee OA, as this joint possibly bears more of the whole body weight, and more exposed to traumatic injuries than does the hip joint [3, 36, 37]. On the other hand, the interest in studying knee OA more frequently compared to hip OA may indeed be explained by the fact that knee OA seems to be more disabling than hip OA, especially for walking [38].

In this review, the mean age of 64 (6) years and the mean BMI of 28.2 (4.5) highlighted that OA is mainly a pathology affecting older and overweight adults. This may possibly be explained by the fact that the joint cartilage degenerative process could be age-related, given that most old people are less or not physically active, and therefore can become overweight, as this may be associated with inactivity [39]. Moreover, it has been reported that older adults spend approximately 60% - 70% of their waking hours engaging in sedentary activities, which significantly increases their risk of functional declines and other negative health outcomes, such as chronic disease development, premature mortality, and obesity [39]. Therefore, it has been suggested that physical exercises must be prescribed in association with patient education and weight loss (if BMI > 25); this will help boost recovery from pain and disability by losing weight and improving exercise compliance [40].

Considering the clinical efficacy of supervised exercises, no statistical significance was found in terms of pain and disability reduction when compared to passive MSKTs. On the other side, HBE improved significantly pain compared to supervised exercises, but the 2 interventions displayed almost equivalent effects in reducing disability. Previous studies concluded that both supervised exercises and HBE are effective in reducing pain severity for people with hip or knee OA [41–43]. It has been previously reported that the effects are maximal around 2 months and after then slowly decrease, becoming no better than usual care at a longer follow-up ranging from 9 to 18 months, and patients with younger age (< 60 years old) not awaiting joint replacement may benefit more from exercise therapy [42]. Although the mechanisms through which exercise may reduce pain severity remain unclear, it is suggested that being physically active lessens the degree of biomechanical change, thereby decreasing the load on joints, increasing joint stabilization, and contributing to better segmental motion [44]. From a more general standpoint, exercise may help reverse muscle imbalance or initiate a pain desensitization process, resulting in an increased pain detection threshold [45–47].

Supervised exercises were found to be less effective than HBE in terms of pain reduction. On one side, the possible explanation should be that, patients undergoing supervised exercises may perform these exercises as prescribed and demonstrated regardless they may be experiencing pain flare. In HBE, patients can adjust the dosage, and the way to exercise to their comfort and ease, as they are not supervised by their physical therapist who could constrain them to exercise in a way that may increase pain sensitization. Thus, it can imply that physical therapists would be advised to consider HBE in association with patient education to help boost exercise adherence. This kind of exercise program should be implemented, as it can be performed either individually or in a group. The 2 modalities can be timely and financially-cost effective.

### Study limitations

The limitations of this review must be mentioned and discussed before discussing the implication for rehabilitation. First, only a limited number of studies (3-5 per outcome) were considered in the different analyses, and the findings could not be extrapolated onto the general population with knee or hip OA since most of the analyzed studies that applied supervised exercises involved patients with different characteristics in terms of sample size, BMI, and durations [22, 23, 28]. Concerning the meta-analysis itself, we only have a limited number of studies included in the different analyses and therefore these analyses may suffer from low statistical power and the results of their results should be interpreted carefully.

Furthermore, the quality of the majority of the included studies [24, 25, 28–30, 33, 34] evaluating the effects of supervised exercises exhibited a PEDro score of moderate quality ranging from 4 to 6/10. This could have increased the risk of bias, and therefore impacted by downgrading the level of evidence of this review. In addition, all of the included studies failed to blind the participants and therapists – which is, however, a well-known limitation of studies in the rehabilitation field.

Regarding study characteristics, supervised exercises slightly differed in their design. Thus, supervised exercises included strengthening exercises to the muscles around the knee, the trunk, the hip, and the ankle, proprioceptive training exercises, static bike, stretch, and open and closed-kinetic chains, and stabilization or neuromuscular control exercises. All these interventions were targeted at recruiting knee, and pelvic girdle muscles for different durations which varied considerably, ranging from 2 weeks to 16 months. This heterogeneity in intervention duration may have impacted the results. It may indeed be suspected that there is a dose-response relationship between the total amount of exercises and the clinical improvement [48]. This highlights the need for further studies assessing various types of interventions (e.g., exercise duration, and intensity). The lack of subgroup analysis with knee OA and with hip OA is also another important limitation but given the small number of RCTs conducted on hip OA (4/12), it was not interesting to perform such kind of analysis.

Finally, the included studies failed to report on exercise adherence, whereas many of them involved HBE. The evidence clearly suggests that the effectiveness of patients’ exercises depends on their exercise adherence; indeed, about 70% of patients with chronic musculoskeletal pathologies do not regularly engage in prescribed exercise programs [49, 50].

Despite these limitations, this review highlights the benefits of both supervised exercises and HBE in terms of pain reduction and improving function in patients with hip or knee OA. Therefore, given their applicability and ease of use (no material needed), these interventions should be promoted in patients and primary prevention.

Moreover, this review considered only RCTs, which may have increased this review’s level of evidence.

### Implications for rehabilitation

Researchers and clinicians should work together to clinically determine the best effective exercise combination and dosage, which can be necessary to reduce pain and disability in the hip or knee OA and, therefore, postpone surgery. Indeed, a clear ‘call to action’ for exercise therapy in hip or knee OA for primary prevention as well as early detection may greatly improve the current management, as they may be relatively easy to apply and use in daily clinics.

For quick improvement and prevention of recurrence, clinicians should consider the combination of supervised exercises and HBE to manage hip or knee OA. They should be advised to provide education and reassurance about managing potential pain flares and inflammation by modifying exercises and physical activity. Interestingly, as a result of COVID-19, there is an urgent need to shift to HBE either guided by mHealth or not.

To increase patients’ motivation, besides patient education, clinicians should prescribe exercises in the context of games, especially in groups (cost reduction, motivation, group effect, etc.). Indeed, the use of a mobile app or connected watch that include different kind of exercises either in the picture or video format may help boost their motivation, and finally the recovery [50]. In this context, the tools should be used as reminders, such as applications showing how to perform exercises with live feedback, and motivating and fun rehabilitation exercises in computer games. Supervised sessions/exercises should be considered in patients with low exercise adherence or in case of pain avoidance and catastrophizing behavior.

Finally, it should be noted that almost all the included studies were conducted in high-income countries (HIC), except one study that was performed in a middle-income country, India [22]. Therefore, the findings of this review may not be extrapolated to other countries (i.e., the specific organization of the care, health literacy, beliefs and cultural background, etc.). This highlights the need of conducting such research and implementing the findings in low- and middle-income countries, taking into account contextual realities [51]. Indeed, as exercise therapies may be of easy applicability and use (no special material needed), they could be easily contextually adapted and implemented in these countries.

## 5. Conclusion

Supervised exercises were found to be less effective in reducing pain compared to HBE and the 2 interventions displayed almost equivalent effects in terms of disability reduction. When comparing supervised exercises to passive MSKTs, no statistically significant differences were found between the two groups in terms of pain or disability reduction. Based on the results of this review, it seems that the best therapeutic approach would be to combine supervised exercises (group) and HBE to maximize the outcomes of the rehabilitation process. Further RCT researches with exercise adherence assessment included are warranted.

## Data Availability

All data produced in the present work are contained in the manuscript

## Conflict of interest

The authors declare that the research was conducted in the absence of any commercial or financial relationships that could be considered a potential conflict of interest.

## Notes

### Competing Interest Statement

The authors have declared no competing interest.

### Funding Statement

This study did not receive any funding

